# Omics Integration Uncovers Mechanisms Associated with HIV Viral Load and Potential Therapeutic Insights

**DOI:** 10.1101/2025.07.29.25332397

**Authors:** Kyle A. Sullivan, Melyssa S. Minto, Xinyu Zhang, William Carr, Bryan C. Quach, Caryn Willis, Alice Townsend, Peter Kruse, Matthew Lane, Richard Morgan, Ke Xu, Bradley E. Aouizerat, Dana B. Hancock, Daniel A. Jacobson, Eric O. Johnson

**Affiliations:** Computational and Predictive Biology Group, Biosciences Division, Oak Ridge National Laboratory, Oak Ridge, TN; GenOmics and Translational Research Center, RTI International, Research Triangle Park, NC; Department of Psychiatry, Yale School of Medicine, New Haven, CT; Department of Biology, Medgar Evers College, City University of New York, Brooklyn, NY; Bredesen Center for Interdisciplinary Research and Graduate Education, University of Tennessee-Knoxville, Knoxville, TN; Veterans Affairs Connecticut Healthcare System, West Haven, CT; Biomedical Informatics and Data Science, New Haven, CT; Translational Research Center, College of Dentistry, New York University, New York, NY; Fellow Program, RTI International, Research Triangle Park, NC

## Abstract

While antiretroviral therapy (ART) has significantly improved disease prognosis in people with HIV (PWH), understanding the biological mechanisms underlying plasma HIV-1 RNA viral load (VL) can inform additional strategies to slow HIV/AIDS disease progression. Here, we integrated multi-omic datasets and used two machine learning network biology tools (GRIN and MENTOR) to identify biological mechanisms associated with VL across 10 cohorts from multiple omics data sets. We integrated the following gene sets: 3 genes from HIV set point VL GWAS, 258 genes whose expression was associated with set point VL in CD4+ T-cells, 143 genes based on DNA methylation associations with VL, and 8 genes previously known to affect the pharmacokinetics of ART. Using GRIN, we retained 194 VL genes based on their high network interconnectivity. We then used MENTOR to collaboratively interpret subsets of these genes and identified the following biological processes: cell cycle checkpoint pathways associated with non-AIDS defining cancers, oxidative stress, viral replication, and interferon signaling. Using these network tools for multi-omic integration, we present a conceptual model of mechanisms underlying HIV VL, and identify drug repurposing candidates to complement existing ART to enhance treatment response and reduce HIV-related comorbidities.

## Introduction

The development of combination antiretroviral therapy (ART) that suppresses HIV replication to undetectable levels in the blood has dramatically increased the life expectancy of persons with HIV (PWH)^1^. With lifelong adherence, current antiretroviral regimens are capable of slowing the progression of HIV infection by decades^1^. Moreover, maintaining HIV viral suppression by reducing viral load (VL) has reduced the risk of transmission and the incidence of HIV infection^2^. Notably, the ability to lower HIV VL can be negatively impacted by molecular consequences arising from prolonged use of alcohol^3^, injection drug use^4^ and/or cocaine use^5^. This impact on preventing HIV VL suppression can result in disease progression from HIV and impair long-term health outcomes in PWH.

Despite the profound impact of current ART on the suppression of HIV infection, PWH develop age-associated conditions at earlier ages than persons without HIV infection (PWoH), including various types of cancer^6,7^. While current antiretroviral regimens result in potent suppression of HIV viral replication in the periphery, prevention of antiretroviral penetration into the central nervous systems remains sub-optimal even among highly ART-adherent persons^8^. In tandem, the persistence of low-grade inflammation among medication-adherent, virally-suppressed PWH is well-established and due in part to changes in immune cell composition that may impact the efficacy of immune surveillance in relation to oncogenesis^9–11^. Of the cancers that PWH can develop, they are typically split into two categories: AIDS-defining and non AIDS-defining cancers. AIDS-defining cancers are named based on their higher incidence following HIV infection and include aggressive B-cell non-Hodgkin’s lymphoma, Kaposi sarcoma, and cervical cancer^12^. However, there are also higher incidence rates of non AIDS-defining cancers that have been reported in PWH^13^. However, the extent to which increased HIV VL leads to elevated risk of non-AIDS defining cancers is poorly understood.

In this study, we used machine learning network analyses to uncover biological pathways and mechanisms from multi-omic datasets that underlie the relationship between HIV VL and comorbid diseases, such as non-AIDS defining cancers. We began by performing a novel epigenome-wide association study (EWAS) meta-analysis to understand how the DNA methylation (DNAm) state of whole blood was affected in response to HIV VL. We then integrated these genes associated with differential DNAm linked to HIV VL (n=143) with genes previously associated with HIV phenotypes: genes whose RNA expression has been associated with HIV set point VL (n = 259)^14^; genes with variants associated with set point VL (n = 3)^15^; and genes previously known to affect the pharmacokinetics of ART drugs (n = 9)^16^. We then examined the biological relationships among these genes by capturing multiple lines of biological evidence from outside experimental data in a multiplex network, and identified the shortest paths between pairs of our multi-omic genes based on this evidence. We then applied MENTOR^17^ to graph-based embeddings of the multiplex network to define sub-clusters of functionally related genes. These sub-clusters were further assessed to identify the underlying biological mechanisms linking increased HIV VL to other comorbidities in PWH. This approach identifies functional pathways associated with HIV VL and sheds light on comorbid conditions (e.g. cancers) in PWH that may be caused by shared pathways affected by VL^18^ and/or by secondary mechanisms such as opportunistic infections or viral-induced inflammation. Lastly, we identified drugs targeting the multi-omic genes and explored their potential as novel therapeutic interventions to mitigate HIV-associated health risks. This approach found currently approved ART, as expected, and uncovered drugs that have anti-cancer targets that may serve as intriguing drug repurposing candidates combined with existing combinations of ART.

## Results

### Meta-epigenome wide association study (EWAS) using 450K and EPIC arrays

We first conducted an EWAS meta-analysis on HIV VL based on the summary statistics from EWAS in 450K and EPIC array samples from VACS. From this meta-EWAS of VL, we identified 147 unique genes at a stringent threshold of Bonferroni-corrected *p* < 0.01 (**Supplementary Tables 1-3)**.

### Network-based gene set filtering using GRIN and shortest paths connections of multi-omic genes

In addition to identifying new genes associated with HIV VL through EWAS, we integrated existing omics studies of HIV VL and ART from multiple data sources (**Figure 1**). This included differential gene expression from CD4+ T-cells of genes associated with HIV VL in untreated PWH^14^, genes associated with HIV set point VL from whole genome sequencing data^15^, and genes known *a priori* to have effects on the pharmacokinetics of HIV ART drugs^16^. Aggregation of genes across these sources resulted in a set of 382 genes used in downstream network analyses (**Supplementary Table 7**).

**Figure 1.**
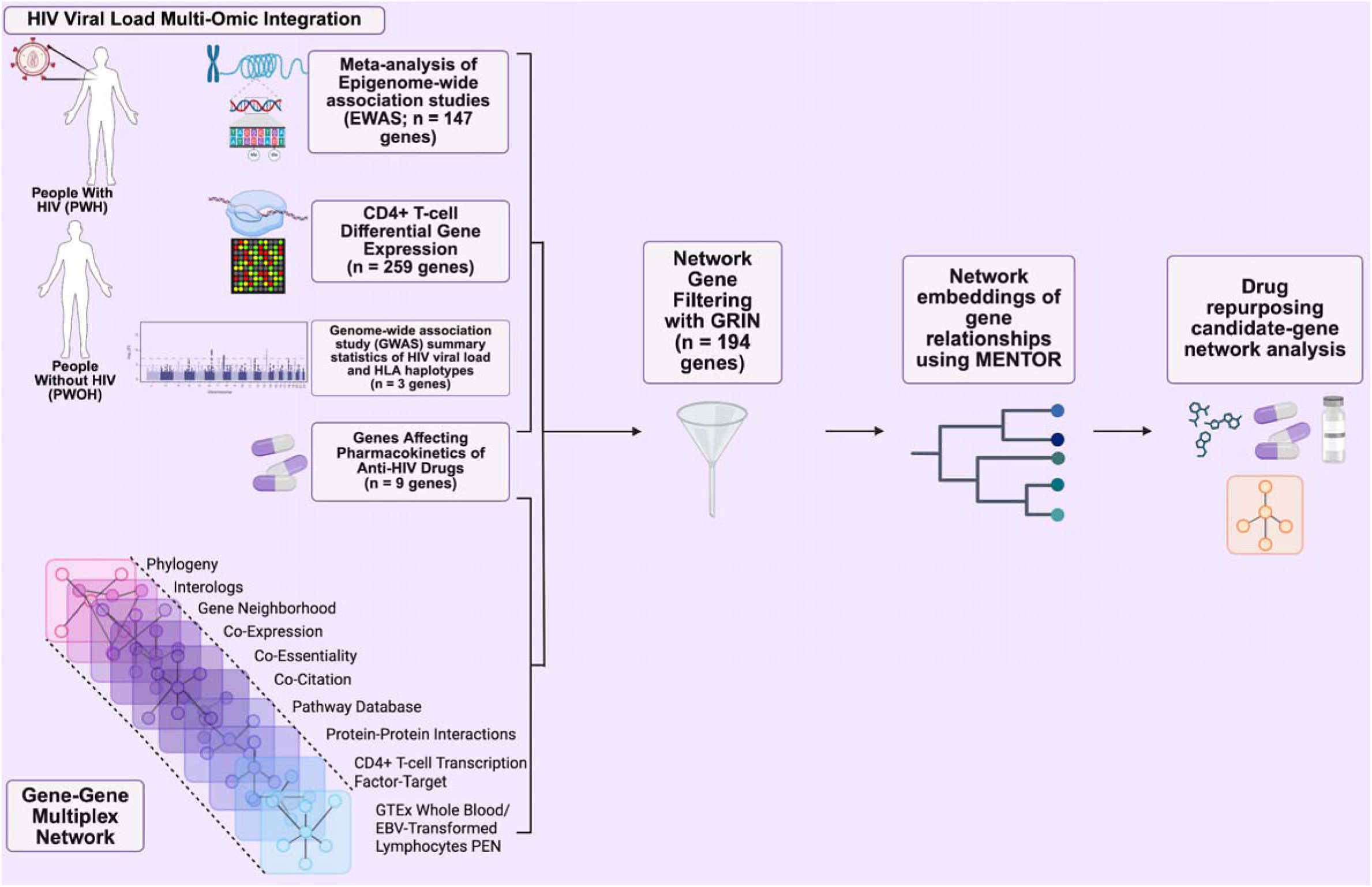
Summary of data sets and workflow for the present study. Genes related to HIV viral load were obtained from multiple omics data sets that were collected from people with HIV and people without HIV. This included a meta-analysis of epigenome-wide association studies (EWAS) to identify DNA methylation sites, differentially expressed genes from CD4+ T-cells, and GWAS of HIV viral load and HLA haplotypes from whole genome sequencing data. These genes were combined with genes known to affect the pharmacokinetics of anti-HIV drugs to form the starting gene set for network-based analyses. Using a 10-layer multiplex network, this starting gene set was first filtered using GRIN software to obtain a set of multi-omic genes, followed by execution of MENTOR software to obtain network-based embeddings. Drug repurposing candidates were then identified from a drug-gene target network.

After identifying 382 genes from these multi-omic sources related to HIV VL and ART, we used GRIN to remove possible false positive genes across the different omics data sets based on identifying highly interconnected genes within a network of gene-gene relationships derived from experimental data sets that were distinct from the multi-omic data sets examined. Coupled with a 10-layer multiplex network containing CD4+ T-cell-specific connections, GRIN refined the starting gene set to 194 highly interconnected genes, which were used in downstream analyses (**Supplementary Tables 8-9**). Using these 194 multi-omic genes, we used shortest paths network analysis to identify specific biological lines of evidence connecting pairs of genes, as opposed to the agglomeration of evidence identified by RWR-based network embeddings as assessed by MENTOR clusters. Out of 194 multi-omic genes, 193 genes were directly connected to at least one other gene in the set based on the topology of the multiplex network, indicating that the GRIN-retained gene set was tightly interconnected (**Figure 2**).

**Figure 2.**
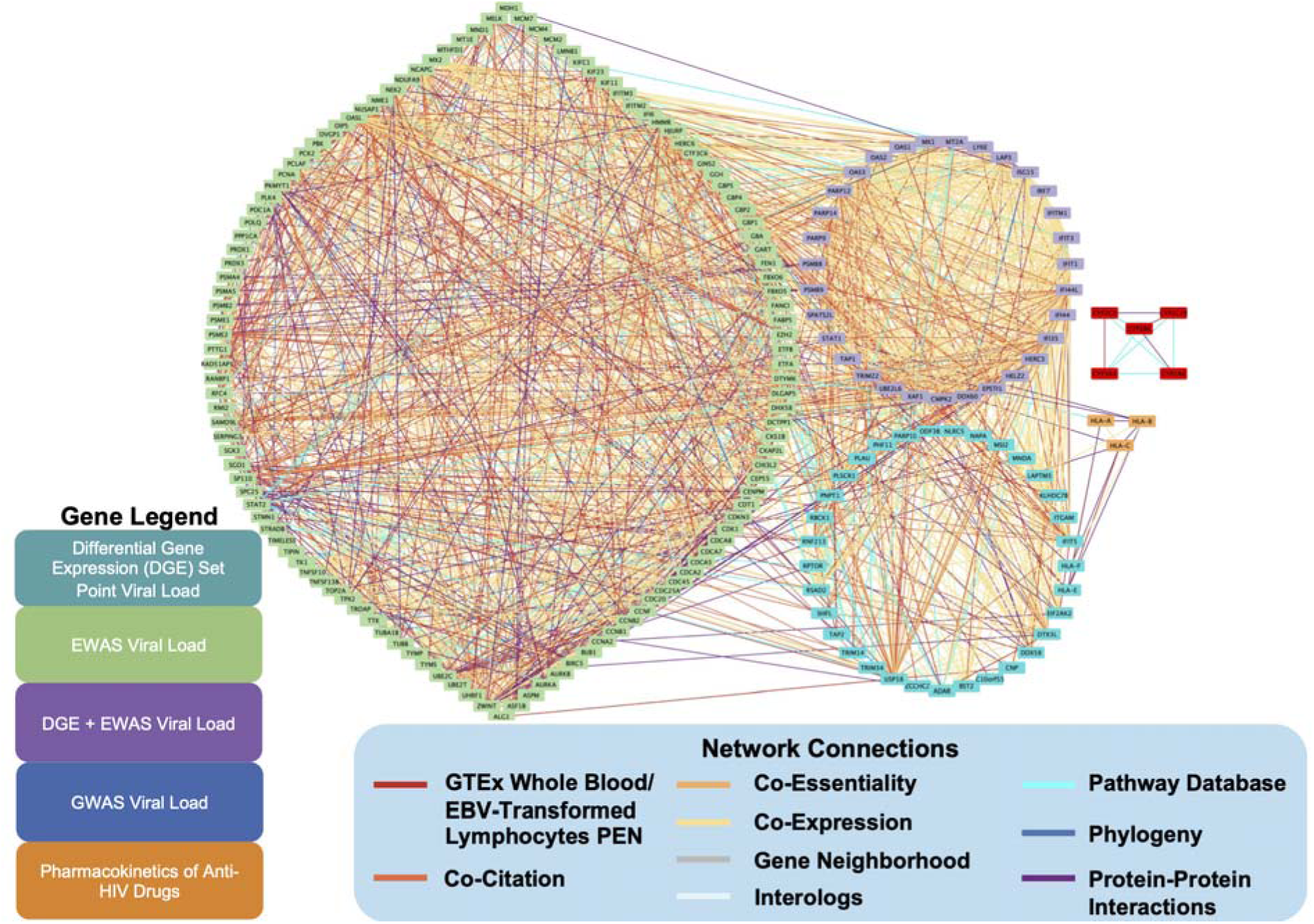
Shortest paths network analysis identifies connectivity among HIV multi-omic genes. Using the 194 GRIN-retained HIV multi-omic genes, shortest paths network analysis was conducted among each of the 194 genes using the network layers present within the multiplex network. By restricting path lengths to direct connections, 193 of the genes were connected to at least one other HIV multi-omic gene directly. Gene legend (color of each node in the network) indicates the data source for each multi-omic gene; network connections legend indicates the type of biological evidence connecting each gene pair.

### MENTOR identifies functionally related gene sets based on network embeddings

To better understand the biological relationships among this network of multi-omic genes, we implemented MENTOR which uses multiple lines of biological evidence, in the form of a multiplex network from normal tissue, to cluster genes into functionally related modules based on network topology. Using the same 10-layer multiplex network used in previous analyses, MENTOR generated 16 modules constrained to a maximum cluster size of 20 genes (**Figure 3**). We confirmed the importance of each line of evidence to the MENTOR clustering, as evidenced by changes to module memberships when individual network layers were ablated from the multiplex (**Supp. Fig 1A**). This was particularly evident in the large differences in the module reassignments observed for *ALG1, GTF3C6, NAPA, PPP1CA*, *PNPT1,* and *GART*, which changed set assignment irrespective of the network layer that was removed (**Supp. Fig 1B**).

**Figure 3.**
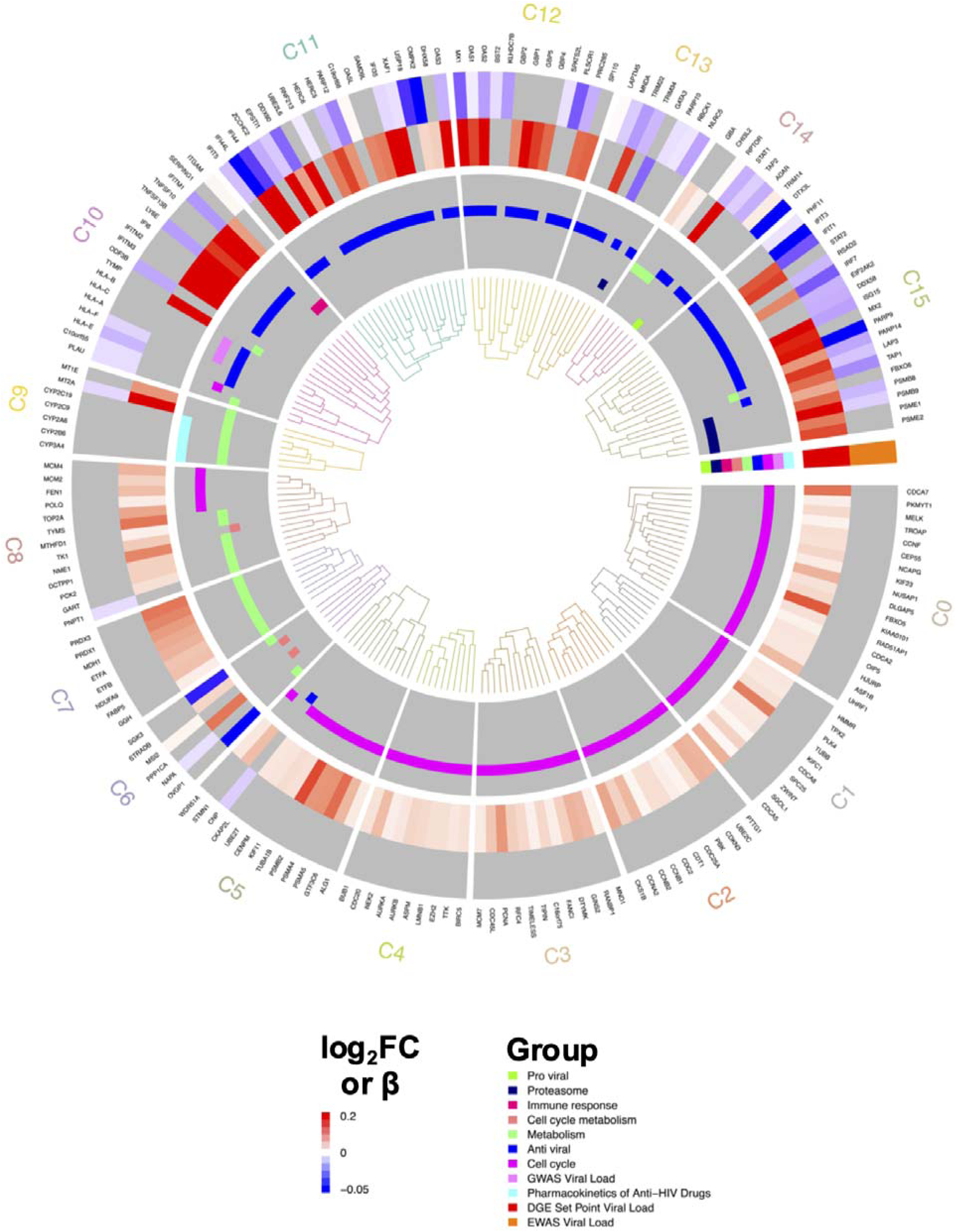
MENTOR network embedding of multi-omic genes and contributions of individual network layers. Applying MENTOR with a 10-layer multiplex network produced a dendrogram of network embeddings from the network relationships. Inner rings of the dendrogram illustrate genes belonging to functional groups (proviral, proteasome, immune response, cell cycle/metabolism, metabolism, and antiviral) and data set (GWAS viral load, pharmacokinetics of anti-HIV drugs, DGE set point viral load, or EWAS viral load). Outer two rings indicate log_2_FC of differential gene expression and β values from EWAS.

Modules were annotated based on the molecular functions of their genes, resulting in three emergent biological themes: Antiviral response (n = 6 modules), metabolism (n = 3 modules), and mitotic cell cycle/cancer (n = 7 modules). Below, we further examine modules linked to these themes to elucidate molecular mechanisms associated with HIV VL, which we integrated into a conceptual model of biological pathways involved (**Figure 4**).

**Figure 4.**
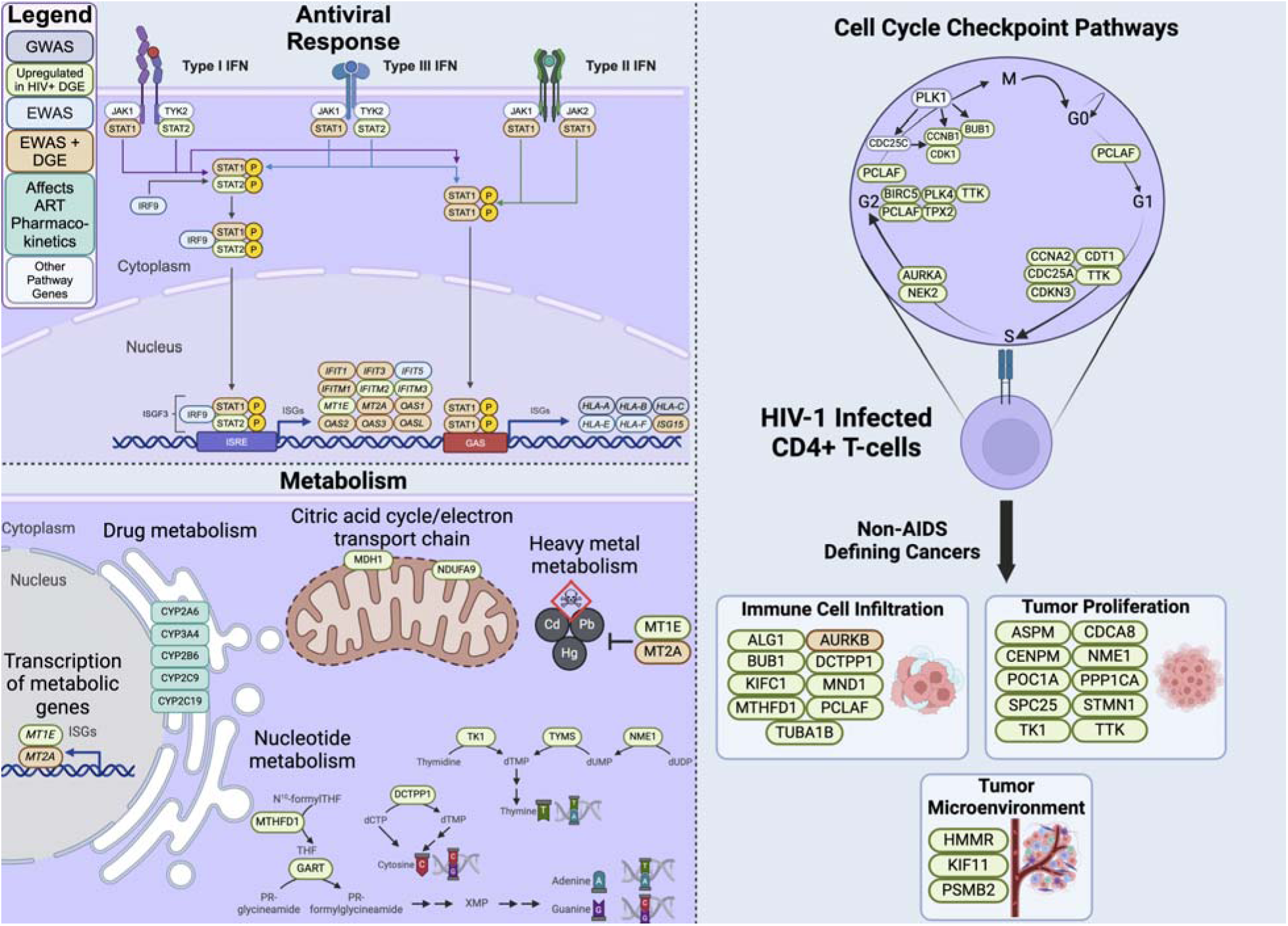
Conceptual model of HIV multi-omic genes. Numerous antiviral response genes from type I, II, and III interferon (IFN) signaling pathways were identified from EWAS and/or differential gene expression (DGE). In addition to the drug metabolism functions of 5 cytochrome P450 genes that are known to affect HIV pharmacokinetics, 8 genes related to nucleotide and heavy metal metabolism were identified via differential gene expression or EWAS, as well as 2 genes (*MDH1* and *NDUFA9*) that contribute to energy production via the citric acid cycle and electron transport chain. Furthermore, 18 genes were identified from EWAS or differential gene expression from CD4+ T-cells have known roles in controlling cell cycle checkpoint, and 22 genes have been implicated in non-AIDS defining cancer-related contexts, including immune cell infiltration, tumor proliferation, and the tumor microenvironment.

#### Antiviral Response

The 6 antiviral response modules (modules 10-15) included functions in antigen presentation, interferon (IFN) signaling, inhibition of HIV replication, and pathogen associated molecular pattern (PAMP) recognition signaling (i.e., RIG-I). Following GRIN filtering and MENTOR clustering, module 11 included *OAS3, OASL, DDX60, DHX58, EPSTI1,* and *IFI35* as functionally related genes in the RIG-I signaling pathway, which is central in initiating the immune response and detecting viral RNA in cells ^19–22^. Our analysis shows that for subjects with higher HIV VL levels, the following genes had significantly upregulated gene expression: *OAS3* [log_2_FC = 0.21], *OASL* [log_2_FC = 0.12], *DHX58* [log_2_FC = 0.05], *IFI35* [log_2_FC = 0.20], *EPSTI1* [log_2_FC = 0.33], *DDX60* [log_2_FC = 0.10]. Additionally, CpG probes near *OAS3*, *EPSTI1*, *IFI35*, and *DDX60* were also statistically significantly hypomethylated. *OAS3*, *OASL*, and *DDX60* work to activate RIG-I signaling, thus promoting type I IFN production to control viral replication ^20,22,21^. Unlike the other significant genes in module 11, *IFI35* has been previously described to inhibit activation of RIG-I^23^. The convergence of these genes onto the RIG-I signalling pathway suggests its importance in antiviral response for patients with high VL. As a submodule within module 11, *OASL*, *IFI35*, *SAMD9L* [log_2_FC = 0.13], and *XAF1* [log_2_FC = 0.28] were most closely associated functionally. As a transcriptional coactivator of IRF-1, human X-linked inhibitor of apoptosis protein-associated factor 1 (*XAF1*) is a pro-apoptotic tumor suppressor that forms a positive feedback loop with IRF-1 to enhance stress-induced apoptosis by stabilizing and activating IRF-^24^. In contrast, *SAMD9L* directly binds to HIV RNA to inhibit translation to mediate antiviral activity^25^.

In addition to identifying functionally related components in the RIG-I signaling pathway that converge on STAT1 activation, this approach also revealed functional associations among components of the ISGylation signaling pathway, which converges on STAT1 activation. In module 11, *HERC5*, *UBE2L6*, *SAMD9L*, *USP18*, and *RNF213* are genes involved in or related to the ISGylation signaling pathway. With the activation of the JAK-STAT signaling pathway, STAT1, STAT2, and methylated IRF9 form a complex that is then demethylated for migration to the nucleus. This complex then binds to ISRE to up-regulate transcription of various interferon-stimulated genes (ISGs), including *UBE2L6, HERC5, HERC6, RNF213,* and *SAMD9L*. Within module 11, *UBE2L6, HERC5, HERC6, and RNF213* were closely associated topologically as a subcluster. For patients with higher levels of VL, *HERC5* [log_2_FC = 0.18], *HERC6* [log_2_FC = 0.13], *UBE2L6* [log_2_FC = 0.16], and *SAMD9L* [log_2_FC = 0.13] were statistically significantly up-regulated from the Rotger et al. differential gene expression study^14^, but only *HERC5* and *USP18* were statistically significant within the meta-EWAS. Their functional association in antiviral innate immune responses had been documented previously to inhibit viral stability^26^, facilitate viral replication^27^, regulating RIG-I autophagy^28^, and acting as an antimicrobial effector^29^. Topologically, these genes cluster most closely with *DDX60* and *EPST11*, genes that promote RIG-I signaling and type I IFN production upstream of ISG gene transcription.

As a separate subcluster within module 11, two genes, *C19orf66/SHFL* and *PARP12*, were identified as functionally associated with higher levels of HIV VL. Both genes have been previously described as ISGs that limit RNA translation by altering ribosomal frameshifting^30,31^ or by directly binding RNA for mRNA translational arrest^32^.

Within two of the antiviral signaling modules (modules 13 and 15), we found genes involved in the ubiquitin-proteasome system (UPS), which is known to be manipulated by HIV proteins to degrade host cellular proteins that repress viral replication and evade host immune response^33,34^. The protein degradation modules included functions in protein degradation (*RBCK1*), metabolism (*LAP3*), and the ubiquitin proteasome (*PSMB8*, *PSMB9*, *PSME1*, and *PSME2*). *LAP3*, which is known to have peptidase activity, was both upregulated with increased VL (DGE log_2_FC = 0.184) but was negatively associated in the meta-EWAS (β = −0.012485). We found *RBCK1*, a transcription factor with ubiquitin transferase activity that has been previously upregulated in ART-naive PWH^35^ and negatively associated in the meta-EWAS analysis (β = −0.005285). Lastly, we found a group of proteasome subunit genes with increased expression with increased VL: *PSMB8* (DGE log_2_FC = 0.152, EWAS β = −0.014965), *PSMB9* (DGE log_2_FC = 0.201, meta-EWAS β = −0.010005) *PSME1* (log_2_FC = 0.131), and *PSME2* (log_2_FC = 0.131). The upregulation of these genes associated with the UPS and their connection to increased VL underscores the importance of the UPS in HIV replication and immune evasion.

#### Metabolism

Our analysis revealed 3 modules (modules 7-9), consisting of 22 genes, that were enriched for various types of metabolism. All metabolism genes were significantly up-regulated in high VL. Genes annotated to metabolism modules included cytochrome P450 genes involved in metabolism of drugs (*CYP2A6*, *CYP2B6*, *CYP2C19*, *CYP2C9*, *CYP3A4*), heavy metals (*MT1E*, *MT2A*) and nucleotides (*DCTPP1*, *GART*, *MTHFD1*, *NME1*, *TK1*, *TYMS*). Furthermore, the proteins encoded by *MDH1* and *NDUFA9* are important in energy metabolism based on their respective roles in the citric acid cycle and electron transport chain.

Of the genes included based on their known anti-HIV drug effects (**Supplementary Table 2**), only the cytochrome P450 genes were retained by the GRIN analysis and formed a subcluster in module 9 using MENTOR. The other subcluster in module 9 consists of interferon stimulated genes, *MT1E* and *MT2A* ^36^, which function in heavy metal detoxification and homeostatic control of intracellular metal ions thus reducing the bioavailability of metals and potentially reducing oxidative stress response in cells ^37,38^. Expression of both MT1E (DGE FDR = 0.004, log_2_FC = 0.11) and MT2A (DGE FDR = 0.004, log_2_FC = 0.25) are positively associated with HIV VL. This is consistent with reports of higher concentrations of lead, cadmium, and mercury ^39–41^ as well as deficiencies of copper, iron and zinc in PWH ^37,41^. The increase of heavy metals can contribute to oxidative stress and inflammation, both of which are exacerbated by HIV infection. A high VL has been associated with heavy metal stress and our data reveals molecular mechanisms that may respond to these cellular environments with the upregulation of MT1E and MT2A.

A subset of these genes have specific functions in nucleotide metabolism, which is a crucial function during the cell cycle S phase when DNA synthesis occurs. These genes can mostly be found in module 8, including *DCTPP1*, *GART*, *MTHFD1*, *NME1*, *TK1*, and *TYMS* ^42^. These findings advance our understanding of the specific genes related to DNA synthesis and metabolism that may be co-opted during early stages of HIV replication such as viral integration and transcription.

#### Cell Cycle/Cancer

The last group of modules discussed, C0 - C6, contained genes with known cell cycle functions. Not only did this include all four phases of the cell cycle, but also mediating G1 to S and G2 to M transitions, and general molecular functions important in the cell cycle including ATP synthesis, microtubule regulation, and oxidative stress. Of the 80 genes in this category, 75 were upregulated with increased VL. Many of these genes have roles in cancer proliferation, specifically in regulating immune cell infiltration (*ALG1*^43^, *AURKB*^44^, *BUB1*^45^, *CCNA2*^46^, *DCTPP1*^47^, *KIFC1*^48^, *MND1*^49^, *MTHFD1*^50^, *PCLAF*^51,52^, and *TUBA1B*^43,53^), tumor microenvironment (*HMMR*^54^, *KIF11*^55^, and *PSMB2*^56^), and tumor proliferation & invasion (*ASPM*^57^, *CDCA8*^58^, *CENPM*^59^, *NME1*^60^, *POC1A*^61^, *PPP1CA*^62^, *SPC25*^63^, *STMN1*^64^, *TK1*^65^, and *TTK*^66^).

Only 3 of these genes were EWAS hits: We observed a negative β for both the HIV-1 assembly inhibitors *CNP*^67^ and *NAPA*^68^ implying hypomethylation at these sites at a high VL. The last EWAS hit, *MSI2*, has functions in cell cycle regulation and is associated with progression and poor prognosis of multiple cancers^69^. One study has shown that *MSI2* is involved in promoting immune infiltration via mediating post translational modifications to a strong driver of inflammation, *HMGB1*^70^. These findings shed light on a potential shared mechanism upregulated with increased HIV VL and the modulation of the immune response of cancer proliferation (**Table 1**).

**Table 1:**
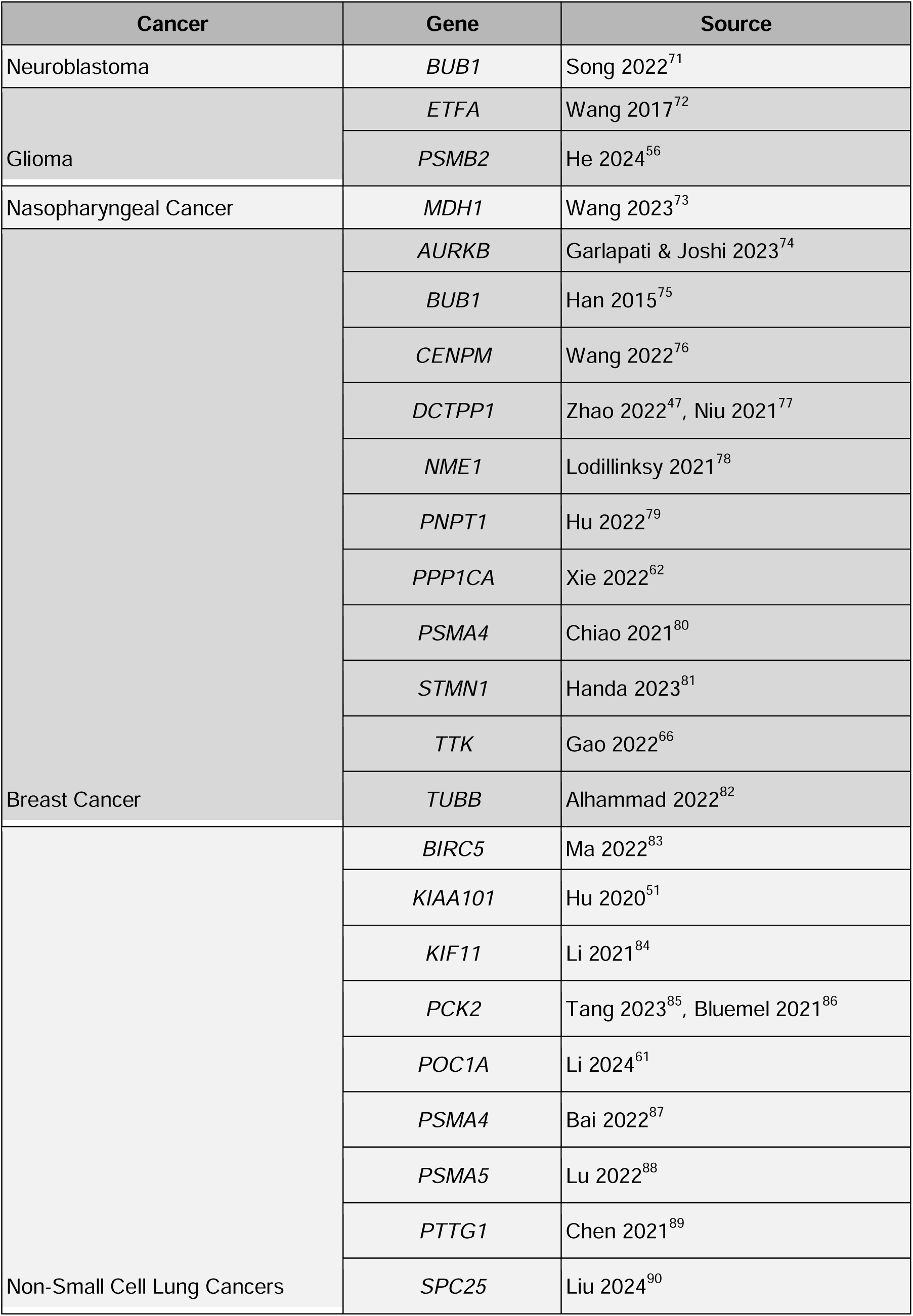
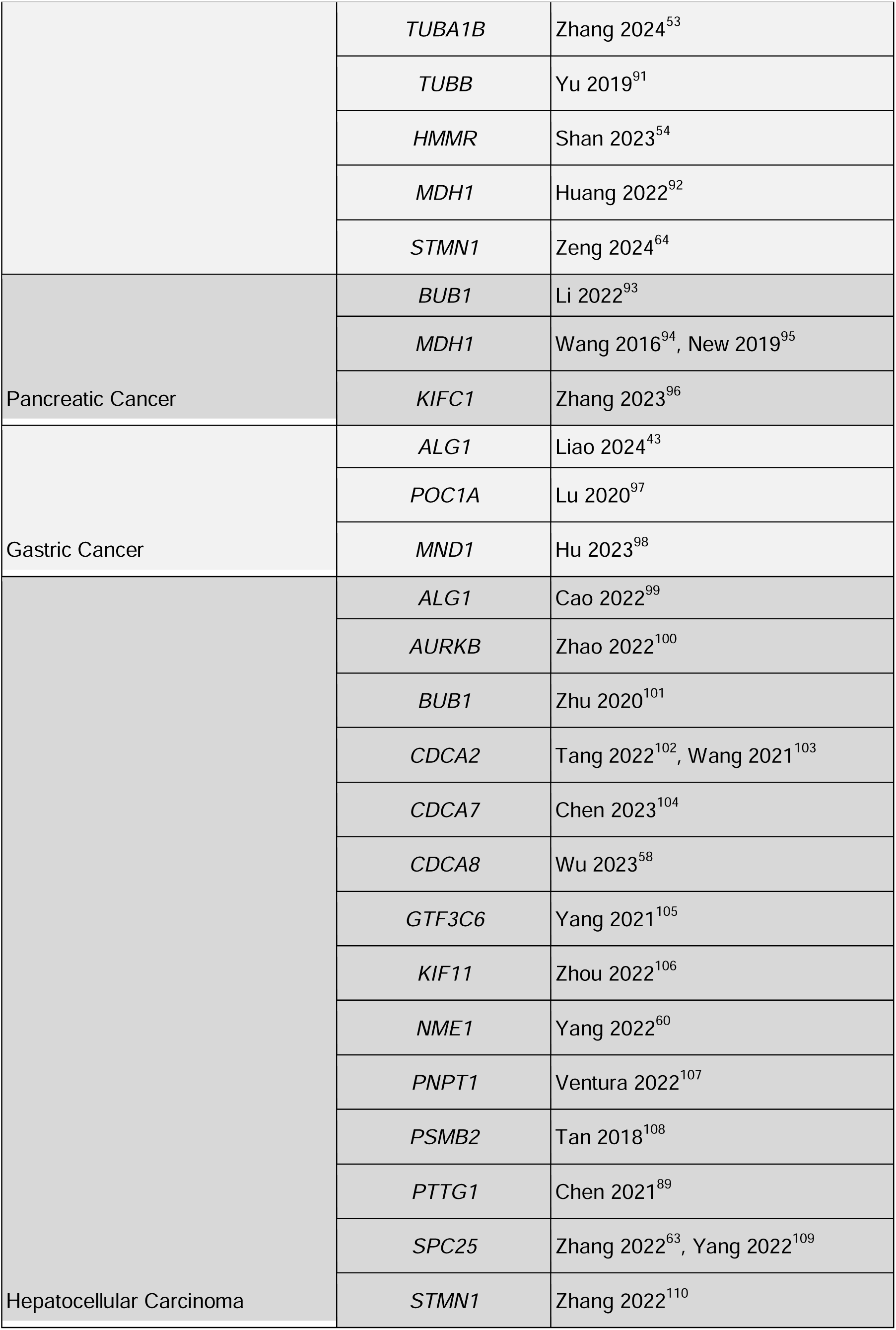
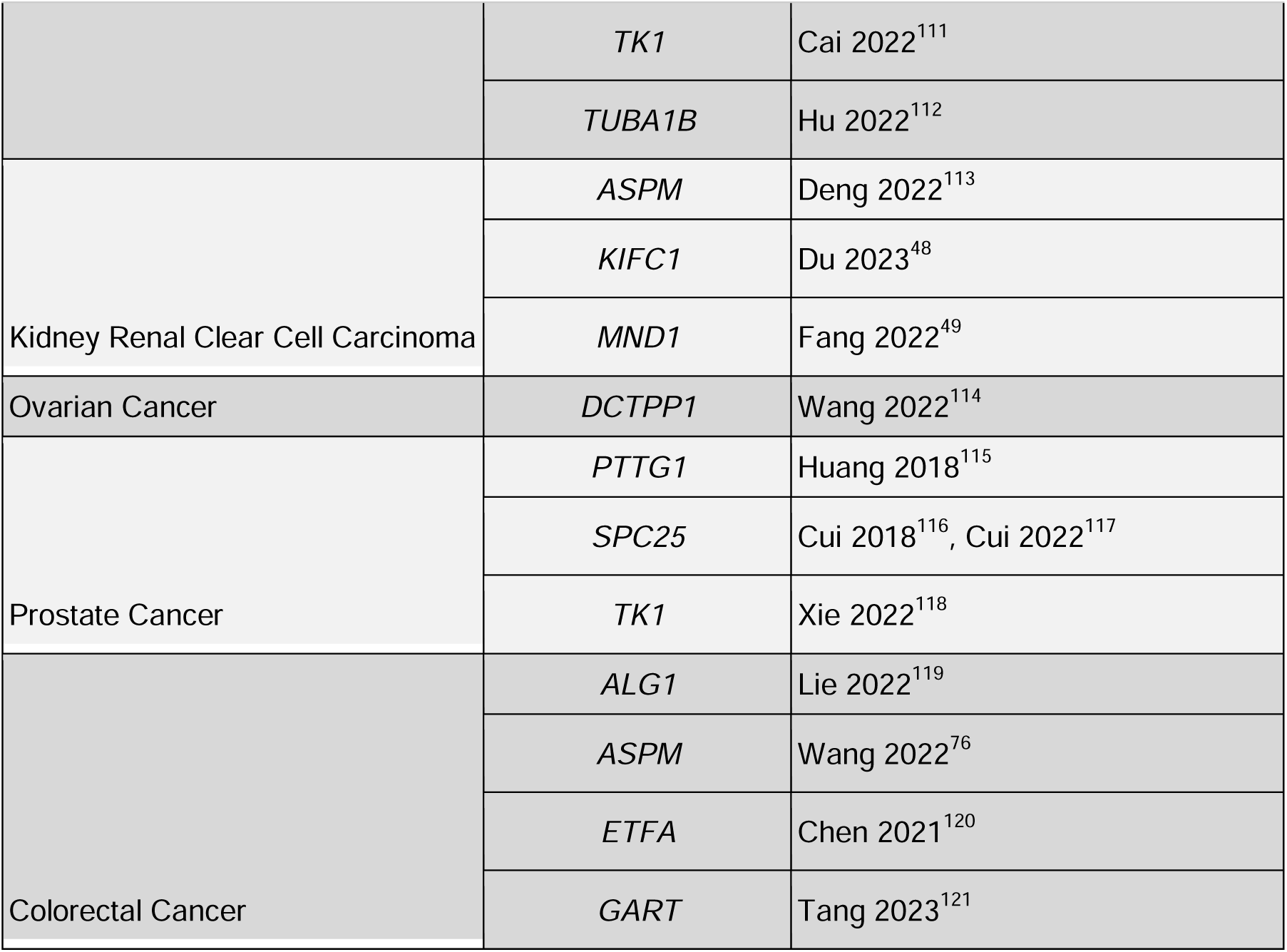
Genes implicated in Non-AIDS defining cancers.

### Drug repurposing candidates through network analysis

After identifying relevant molecular functions and processes among the GRIN-retained multi-omic genes, we sought to identify which genes were known to be druggable targets. Identifying druggable genes would help prioritize candidates for drug repurposing to reduce HIV VL, which could supplement current ART regimens for individuals with lessened treatment response. We used DrugBank to identify a network of drugs that were known to target one of the 194 GRIN-retained genes from our multi-omic gene set (**Figure 5**). As expected, several current approved ART drugs were identified in this set that are known NRTIs: abacavir, lamivudine, tenofovir, and zidovudine (targeting *DYTMK* encoding deoxythymidylate kinase, *HLA-B*, *NME1* encoding NME/NM23 Nucleoside Diphosphate Kinase 1, and *TK1* encoding thymidine kinase 1). The network also included ART from other drug classes (atazanavir, bictegravir, cobicistat, darunavir, efavirenz, elvitegravir, enfuvirtide, fosamprenavir, maraviroc, nevirapine, rilpivirine, ritonavir, saquinavir) targeting cytochrome P450 genes in our set (*CYP2B6*, *CYP2C19*, *CYP2C9*, and *CYP3A4*) that were excluded from the main analysis (**Supplementary Table 11**).

**Figure 5.**
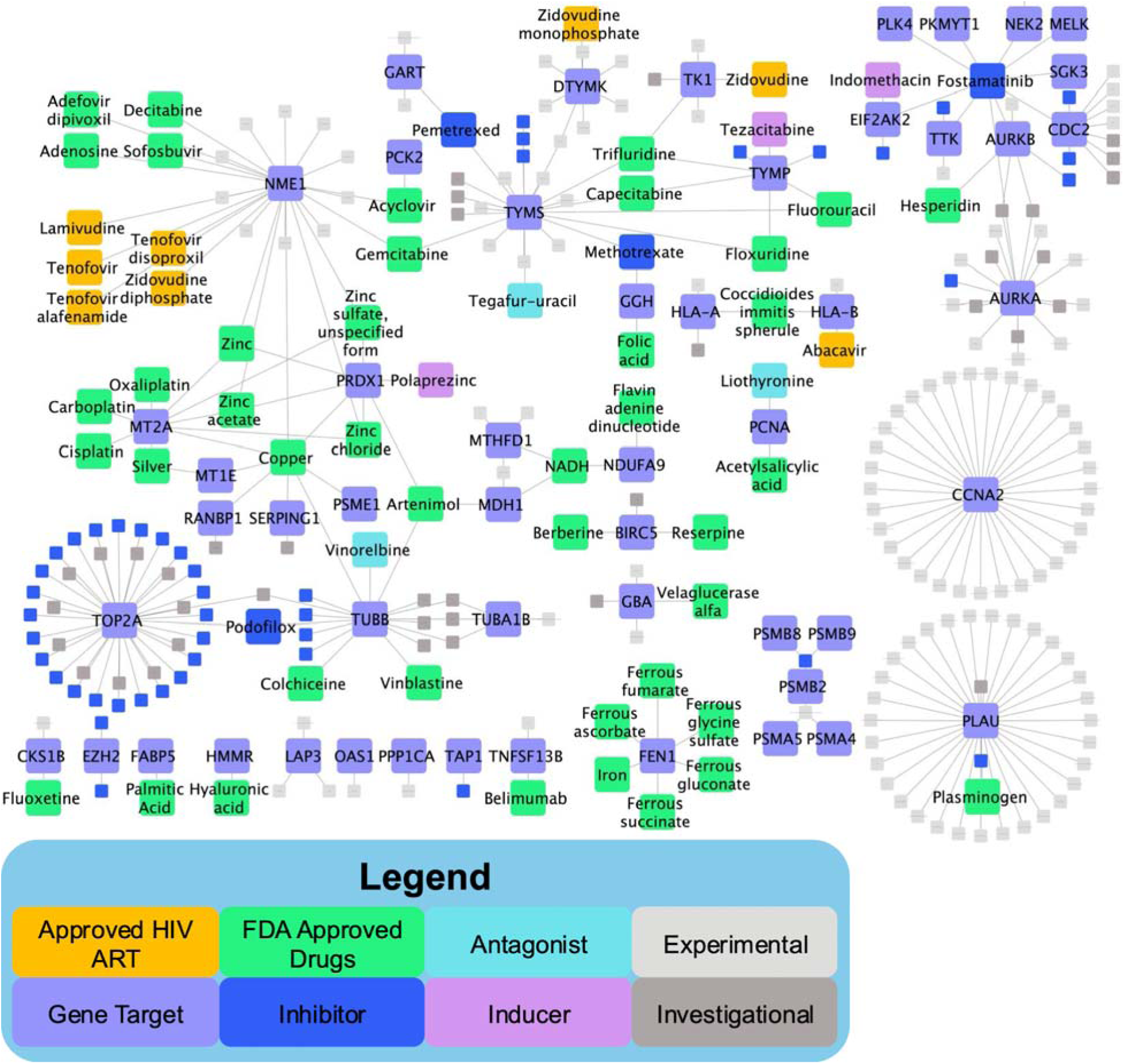
Drug repurposing candidates in drug-gene target network from HIV multi-omic genes. Connecting multi-omic genes (light purple) to drugs known to affect their encoded proteins identified currently approved HIV antiretroviral therapies (orange) in the network, including tenofovir and four other drugs targeting NME1, and three other drugs targeting *DTYMK*, *HLA-B*, and *TK1*. In addition to currently FDA-approved drugs (green), drugs known to inhibit (dark blue) or antagonize (light blue) gene function were identified, while only 3 drugs were identified that induce (light pink) gene function (inducing *EIF2AK2*, *PRDX1*, and *TYMP*). In addition to these drugs with current approval or known mechanism of action, several experimental or investigational drugs were identified as targeting HIV multi-omic genes.

In addition to these approved ART, a number of other drugs targeted multiple proteins encoded by our gene set. These drugs included fostamatinib (an inhibitor of proteins encoded by *AURKA*, *AURKB*, *CDC2*, *CYP3A4*, *EIF2AK2*, *MELK*, *NEK2*, *PKMYT1*, *PLK4*, *SGK3*, and TTK), acyclovir (targeting proteins encoded by *NME1* and *PCK2*), and a number of anticancer drugs such as capecitabine (*CYP2C9*, *TYMP*, and *TYMS*), enzastaurin (*AURKA* and *AURKB*), epothilone D (*CYP3A4*, *TUBA1B*, *TUBB*), fluorouracil (*CYP2A6*, *CYP2C9*, *TYMP*, and *TYMS*), gemcitabine (*NME1* and *TYMS*), methotrexate (targeting proteins encoded by *CYP3A4*, *GGH*, and *TYMS*), and pemetrexed (*GART* and *TYMS*). Further, drugs known to treat other chronic conditions were in our network, such as peginterferon alfa-2b (used to treat hepatitis-induced liver disease; *LEFTY2*) and reserpine (used to treat high blood pressure; *BIRC5*). Together, we observed a substantial overlap in the number of anticancer drugs targeting proteins encoded by our multi-omic HIV gene set.

## Discussion

In the present study, we leveraged network algorithms to identify biological pathways associated with HIV VL and ART from multiple omics types. We performed a novel meta-EWAS to identify genes associated with DNA methylation state in CD4+ T-cells from PWH, which identified 147 genes associated with differentially methylated CpG sites identified by a meta-EWAS. By combining these EWAS-associated genes with HIV set point VL associated gene expression differences in CD4+ T-cells, host genetics associated with HIV set point VL, and genes associated with ART pharmacokinetics, we identified a subset of 194 multi-omics genes that were tightly interconnected within a multiplex network. By identifying important metabolic and cell cycle functions among these genes, we extend beyond antiviral signaling pathways as targets in combating elevated HIV VL. Further, by identifying drug repurposing candidates based on these GRIN-filtered multi-omic genes, new candidates for drug repurposing were highlighted for future experimental follow-up. These candidates may be included with current ART drug combinations to maintain lower HIV VL or suppression, which can promote increased longevity or quality of life in PWH.

Using GRIN and MENTOR, we identified genes associated with antiviral signaling pathways across 6 mechanistic modules. These antiviral-associated genes both recapitulated known host response pathways following HIV-1 infection, as well as genes associated with host response to other viruses that had not been previously characterized in response to HIV-1 infection. Of genes known to be associated with HIV-1 infection, MENTOR module 11 contained *DDX60*, and HIV-1 viral protein R (VpR) can promote *DDX60* expression in the host to enhance its replication^122,123^. Furthermore, the DDX60-like protein (*DDX60L*) is a host factor required for HIV replication^124^. Additionally, HIV tat protein administration *in vitro* has been previously described to induce its expression along with several other genes identified in this study, including *PARP9* (module 15) and *IFITM1* (module 10)^125^. However, there are also instances of multi-omic genes having important roles in antiviral signaling that have not been previously characterized in the context of HIV-1 infection. For example, Oshiumi et al. showed that a knockout of *DDX60* reduces the activation of RIG-I signaling and significantly reduces virus-induced type 1 IFN production in response to herpes simplex virus and vesicular stomatitis virus^21,22^. Similarly, *DHX58*, which is a non-signaling member of RIG-I like receptors, also activates innate immune responses to RNA viruses^126^; however when *DHX58* is methylated, it dampens antiviral immunity and enhances RNA viral infection^127^. In addition, in the RIG-I signaling pathway, epithelial-stromal interaction 1 (*EPSTI1*) has been previously described to promote STAT1 and NFkB nuclear translocation and activation by degrading IkB as well as promoting proinflammatory M1 macrophage polarization in mice^128^.

We also found that genes linked to increased VL were associated with increased expression of genes related to drug, heavy metal, and nucleotide metabolism. These results support known HIV biology, particularly regarding anti-HIV drug metabolism through cytochrome P450 genes and the dysregulation of heavy metal metabolism involving *MT1E* and *MT2A*. Additionally, a subset of nucleotide metabolism genes showed significant disruption in relation to VL. HIV is known to manipulate host cellular machinery to create favorable conditions for its replication, including the upregulation of transcriptional pathways that enhance HIV mRNA synthesis^129^. This metabolic reprogramming is not unique to HIV infection but is also observed in aggressive cancer phenotypes, where pathways involved in glycolysis, lipogenesis, and nucleotide synthesis are similarly altered to support rapid cellular proliferation and survival ^130–132^.The parallels between HIV-driven metabolic shifts and those seen in cancer suggest potential shared mechanisms that could be further explored for therapeutic targeting.

While the introduction of ART has reduced the overall cancer risk in PWH, cancer remains a critical health concern, with risks varying across cancer types. AIDS defining cancers, which include Kaposi Sarcoma (KS), non-Hodgkin Lymphoma (NHL), and Invasive Cervical Cancer (ICC), tend to be inversely correlated with CD4+ T-cell count^133,134^. Notably, PWH have a 11-17-fold greater risk in developing NHL compared to the general population, which has decreased from 25- to 150-fold greater risk with the prevalence of ART^135^. This is consistent with reports of the association of HIV-1 viral suppression with lower incidence of both AIDS-defining and non-AIDS defining cancers^18^. Despite ART’s success in decreasing AIDS-defining cancers, non-AIDS-defining cancers have emerged as greater risk in PWH with a two-fold greater incidence compared to AIDS-defining cancers^133^. Non-AIDS defining cancers can be broken down into virus-related and non-virus related. Virus-related cancers that remain an increased risk to PWH include Hepatitis B and C-associated hepatocellular carcinoma, HPV-related oropharyngeal and anal cancers^133^, EBV and HPV-related AIDS-Burkitt lymphoma^135,136^, and HHV8 related Primary Effusion Lymphoma^135^. Similarly, non-virus related cancers that remain at increased risk to PWH include melanoma and esophageal cancers^136^. Studies have proposed that chronic immunosuppression contributes to heightened cancer risk in PWH^137^. However, viral and non-viral cancers still at an elevated risk after the introduction of ART suggest that there are other oncogenic mechanisms that may play a role. Both behavioral and molecular mechanisms have been shown to contribute to cancer risk. Intravenous drug use and smoking have both been associated with increased cancer risk for PWH^138^. At the molecular level, immunodeficiency/low CD4+ T-cell count has been shown to upregulate inflammatory mediators such as IL6^139^ and be associated with increased incidence of viral-related cancers^140^. The HIV virus itself has pro-oncogenic effects as it has been shown that HIV-1 viral protein R alters DNA repair pathways (Okuma et al, 2020) and HIV-1 matrix protein p17 promotes the activation of EBV latent oncoprotein (LMP-1)^133,139^. Lastly, there are several ties to HIV and the promotion of angiogenesis, which is essential for the growth and metastasis of cancer. HIV has been shown to lead to the differential splicing of transcripts that regulate angiogenesis (i.e. VEGF, proliferin, IL2, IL18, FGF, HIF1a)^141^ and the upregulation of IL6 which increases the expression of the angiogenesis factor PTTG1^139^. Both HIV and cancer share mechanisms in immune evasion to promote cell proliferation. Oxidative stress and redox imbalance creates an environment conducive to unchecked replication^142^. Interestingly, a therapy which targets the immune checkpoint molecule PD-1, Pembrolizumab, has been shown to not only inhibit tumor growth but also reduce HIV latent reservoir^143^. This highlights the opportunity for repurposing cancer therapies that target immune checkpoint pathways for HIV treatment.

Using DrugBank^144^, we identified several drugs targeting multiple HIV VL genes including anticancer drugs and currently-approved ART. This included 7 anticancer drugs targeting 14 of the genes in our set: capecitabine, enzastaurin, epothilone D, fluorouracil, gemcitabine, methotrexate, and pemetrexed. Previous work has demonstrated that some highly active ART also possess anticancer activity^145^. Thus, future work is merited to explore the extent to which these 7 anticancer drugs could be repurposed as ART, given that they target genes associated with HIV VL. Notably, fostamatinib, a phosphodiesterase inhibitor, targeted 10 of our genes. To our knowledge, fostamatinib has not been tested for efficacy as a combination therapy with ART drugs for the purposes of treating HIV. Fostamatinib was tested in clinical trials for efficacy against COVID-19 infection, and was moderately effective at reducing 28-day mortality rates in patients with severe or critical COVID-19 disease^146^. Future studies are warranted to test the efficacy of fostamatinib and other drug compounds in the drug-gene target network for their capacity to reduce HIV-1 VL. While we included cytochrome P450 genes (*CYP2A6*, *CYP2B5*, *CYP2C9*, *CYP2C29*, and *CYP3A4*) that were known *a priori* to affect the pharmacokinetics of ART, for drug repurposing analyses we excluded these drugs. The approved ART targeting non-cytochrome P450 genes were largely NRTIs: abacavir (targeting HLA-B); lamivudine and tenofovir (targeting *NME1*) and zidovudine (targeting *NME1*, *DTYMK*, and *TK1*) ^147^. Intriguingly, other antiviral drugs also appeared in this drug-gene target network such as acyclovir and sofosbuvir, targeting *NME1*. While acyclovir is typically prescribed as an antiviral for herpes simplex virus, previous reports indicate that acyclovir reduces HIV mRNA levels in the plasma of PWH compared to patients receiving placebo drugs^148^. While sofosbuvir has not been clinically proven to lower HIV VL^149^, the *NME1* gene that it targets is targeted by five approved ART and acyclovir. This supports the notion that our drug-gene target network analysis may identify additional useful candidate drugs for repurposing to reduce HIV VL. Further, gemcitabine (targeting *NME1* and *TYMS* genes in the network) reduced HIV-associated disease progression in a mouse model of HIV-1^150^, indicating its possible clinical utility. Finally, it is important to note that some of the drugs in this network act as antagonists of target genes while other drugs are agonists of the genes. Thus, when the mechanism of action is known, it is important to consider whether drugs that inhibit gene function are targeting genes that exhibit up- or down-regulated gene expression or are associated with hypo or hypermethylated regions of DNA. These considerations will help accelerate prioritization of drug repurposing candidates by taking into consideration the known effects on gene function associated with HIV-1 VL.

While this study provides valuable insights, there are a few limitations that should be acknowledged. One limitation is the reliance on microarray data for expression and methylation. For expression, this involved multiple probes per gene. For methylation, there could be a one-to-one, many-to-one, or one-to-many CpG probe to gene mappings. To integrate these results, genes linked to multiple CpG probes were assigned an overall effect size by averaging probe-specific effect sizes. While this approach helps in aggregating the data for GRIN and MENTOR, it may obscure details in variations present in the individual probes. Our second limitation arises in the integration of the DGE and EWAS data for network building and analysis. The DGE data were sampled from individuals not on ART while the EWAS data were obtained from individuals on ART. This discrepancy in viral suppression may introduce discordance in how gene expression and methylation are related to HIV.

Our comprehensive analysis workflow, which integrates data from a novel EWAS meta-analysis, transcriptomics, and literature-based evidence, highlights the power of multimodal approaches in elucidating complex biological mechanisms. In addition to corroborating established HIV-associated pathways, including RIG-I signaling and cell cycle disruption, our study’s multilayer network approach revealed shared molecular mechanisms between HIV VL and cancer, particularly in pathways related to immune cell infiltration, tumor microenvironment, and tumor proliferation. These insights underscore the potential for repurposing existing cancer therapeutics for HIV treatment, as demonstrated by our drug repurposing analysis, which identified candidates such as capecitabine, epothilone D, gemcitabine, methotrexate, and pemetrexed. This convergence of HIV and cancer biology offers a novel avenue for therapeutic development and investigation into the translational potential of these findings.

## Methods

### Epigenome-wide association study (EWAS) meta-analysis

The Veteran Aging Cohort Study (VACS) is a nationwide longitudinal cohort of veterans including PWH and PWoH designed to study HIV infection and disease progression ^151^. A total of 528 samples of PWH from the VACS Biomarker Cohort, a subset of the VACS, were included in the analysis. The majority (86%) of the VACS sample were of African American/Black (AA) ancestry, and all samples were collected from male participants. Among PWH, 78.3% of the participants self-reported good adherence to their ART regimen. Approximately half of the participants self-reported cigarette smoking, and the majority reported drinking alcohol.

The methylation data from this cohort were previously published and deposited in GSE107080 and GSE107082 in the Gene Expression Omnibus (GEO) repository. Methylation of DNA extracted from whole blood was profiled using the Illumina HumanMethylation 450K Beadchip and EPIC Beadchip. All DNA samples were processed at Yale Center for Genomic Analysis. To reduce batch effects, all samples were randomly placed on each array. Quality control and probe normalization were performed as previously described by Lehne et al. ^152^. We removed 11,648 probes on sex chromosomes and 36,142 probes within 10 base pair single-nucleotide polymorphisms. Samples with a sample call rate < 98% were excluded. The detailed description of data processing and quality control was reported elsewhere ^153^.

We adapted the Control Probe Adjustment and reduction of global CORrelation (CPACOR) pipeline proposed by Lehne et al. ^152^ to provide further quality control and to remove batch effects. CPACOR leverages control probes that are designed in 450K and EPIC arrays to adjust for background noise. Principal Components 1-30 derived from control probes were used in our modeling framework per the CPACOR recommendation. We performed EWAS on VL in 450K and EPIC samples separately using a two-step regression analysis^154^. The first regression model addressed global covariates that may confound the association of methylation with VL. These included age, self-identified ancestry information, ART adherence, smoking, intravenous drug use (IVDU), cell counts, 30 methylation control probe principal components, and six cell type proportions estimated by using the Houseman method^5,155^:

> Model 1): *Quantile normalized β value ∼ log*_10_*(VL) + age + race + ART adherence + IVDU + smoking status + White Blood Cell count (WBC) + 6 cell type proportions Principal Components (PCs) 1 - 30 on intensities of control probe*

We then performed a principal component analysis on the resulting regression residual β values and included the first 5 PCs in the following regression model to further control for unmeasured confounders: (2).

> Model 2):*Quantile normalized β value ∼ log*_10_*(VL) + age + race + ART adherence + IVDU + smoking status + WBC + 6 cell type proportions + PCs 1 - 30 on intensities of control probe + PCs 1 - 5 on residuals from Model 1*

Results from Model 2 for both methylation arrays datasets resulted in low inflation. We conducted an EWAS meta-analysis on VL using the summary statistics from the separate EWAS in the 450K and EPIC samples. We performed inverse-variance weighted meta-analysis as implemented in the METAL program^6^ using effect sizes and p-values for each CpG probe. All CpG sites with false discovery rate < 0.1 in the meta-analysis were included in **Supplementary Table 1**, and CpG sites were assigned to nearest genes using the supplied Illumina manifest annotation file aligned to the GRCh37 genome assembly. Genes associated with CpG sites that had Bonferroni-corrected meta-analysis *p* < 0.01 were used as part of the input gene set for GRIN (see Gene Inclusion; n = 147 genes; **Supplementary Tables 2-3**). For meta-analysis significant CpG probes common between the 450K and EPIC array EWASs (n = 249 probes), a CpG effect size was calculated by averaging the effect sizes from the separate 450K and EPIC array EWASs (**Supplementary Table 2**). Genes with a 1:1 CpG-to-gene mapping were assigned the corresponding CpG probe effect size, resulting in 147 total genes. Genes annotated to multiple significant CpG probes were assigned an effect size by averaging across the corresponding CpG probe effect sizes (**Supplementary Table 3**).

### Gene Inclusion

A total of 382 genes in the input gene set were aggregated from multiple sources for downstream network analyses to elucidate biological pathways associated with HIV VL (**Figure 1**). Genes associated with HIV VL were selected by: 1) the novel meta-EWAS presented in this article (n = 147) (**Supplementary Table 3);** (2) differential gene expression from CD4+ T-cells of PWH pre-ART with a threshold of FDR-adjusted *p* ≤ 0.01 (DGE; n = 259) (**Supplementary Table 4**)^14^; (3) Genome Wide Association (n = 3) from a multi-cohort study^15^ derived from the the International HIV Controllers Study, the AIDS Clinical Trials Group, (**Supplementary Table 5**); and (4) genetic variants known to affect drug metabolism^16^ of ART (n = 9) (**Supplementary Table 6**). Omics were selected from pre-ART cohorts (differential gene expression, some cohorts from Genome Wide Association) in order to capture changes caused by HIV viral replication, as well as other omics (EWAS) in order to integrate molecular changes from PWH who are post-ART to reflect multiple molecular pathways that are affected in this population.

### Multiplex Network Construction

We first built a multiplex network comprised of weighted gene-gene connections for downstream multi-omic integration from 10 different types of biological evidence, with a particular emphasis on CD4+ T-cell-specific connections. From 10 network layers, RWRtoolkit^156^ was used to construct a multiplex network using default parameters (delta = 0.5).

First, we incorporated 7 network layers from HumanNet ^157^: co-citation, co-essentiality, co-expression, molecular pathway databases (e.g., KEGG, Reactome), gene neighborhood, gene functions inferred from protein-protein interactions from orthologs of model organisms, and phylogenetic gene profile relationships. A protein-protein interaction network was also merged from literature-curated connections from HumanNet in addition to protein-protein interactions derived from STRING ^158^ (protein.actions.v11.0, min score = 700, mode=binding).

Next, we curated a CD4+ T-cell-specific transcription factor-target network. Using publicly available chromatin immunoprecipitation (ChIP)-seq data from ChIP-seq Atlas ^159^, we downloaded ChIP-seq BED files for peaks associated with the following 34 transcription factors: BCL6, BRD2, BRD3, BRD4, CREBBP, CTCF, EP300, ETS1, FOS, FOSL1, FOXP3, HDAC1, HDAC2, HDAC3, HDAC6, HMGN1, INO80, IRF4, JUNB, KAT2B, KAT5, KAT8, MYC, NFATC1, NFATC2, NFKB1, PRKCQ, PSIP1, RELA, REST, RUNX1, SRCAP, STAT5A, and YY1.

ChIPseeker ^160^ was used to annotate significant peaks to genes, with significant peaks defined as peaks with FDR-adjusted *p*-value < 1e^-22^. Significant ChIP-seq peaks proximal to the gene were assigned as target genes of the corresponding transcription factor based on ChIPseeker peak annotations to peaks within 1kb of the TSS, peaks within a gene body, or peaks within 300 bp downstream of a gene.

Finally, a blood-specific gene-gene network was constructed using the explainable-AI algorithm iterative random forest leave-one-out prediction (iRF-LOOP ^161^) from publicly available GTEx ^162^ RNA-seq data. Transcript counts from whole blood and Epstein-Barr virus-transformed lymphocytes were input into iRF-LOOP, and edges with an importance score >= 0.05 were included in the final network.

### GRIN gene set filtering

GRIN (Gene set Refinement using Interacting Networks)^163^ was used to reduce potential false positive genes associated with HIV VL using the 10-layer multiplex network and the set of 382 genes aggregated from prior studies as inputs. Briefly, GRIN uses the random walk with restart (RWR) algorithm on the multiplex network within a leave-one-out (among the genes) cross validation scheme, which was used to rank each of the 382 aggregated genes in the primary gene set. These ranks were then compared to a null distribution derived from applying the RWR leave-one-out scheme to 100 random gene sets of equivalent size to the primary gene set, and a Mann-Whitney U test was used to compare the rank distributions of the user’s gene set to the null distribution using a sliding window. The *p*-values were then plotted at each point of the sliding window, and the elbow point of this curve was calculated to identify the threshold at which the ranks from the primary gene set no longer departed from the null distribution ranks. Based on this inflection point, 194 multi-omic genes were retained by GRIN.

### Shortest paths network traversal

Using RWRtoolkit ^156^, a shortest path between each pair of GRIN-retained multi-omic genes was calculated using the same 10 network layers used by GRIN. From all possible pairs of shortest paths calculated, direct gene-gene connections (“one-hop” connections) as well as genes separated by a shared gene (“two-hop” connections) were further studied to understand biological relationships between gene pairs. This one- and two-hop shortest paths network was visualized using Cytoscape ^164^ (version 3.10.0).

### Network-based gene embeddings using MENTOR

The MENTOR (Multiplex Embedding of Networks for Team-Based Omics Research ^17^) algorithm partitions a gene set into functionally related gene groups, known as modules which facilitates the efficient distribution and interpretation of gene sets among research teams. The input used for MENTOR was the 194 multi-omic genes and the 10-layer multiplex network constructed for GRIN. Briefly, using RWR, MENTOR traverses the multiplex network, starting from an initial gene from the input gene set, called the seed gene. This process was repeated with each gene in the input set considered as the seed gene, resulting in a score vector for each gene within the multiplex. The score vector for a given multiplex gene consists of probabilities of the random walkers reaching the given gene when starting from the seed genes (i.e., the multi-omic genes). The mean scores-vs-ranks curve was utilized to filter the score vectors to the most strongly connected genes within the network. Subsequently, the pairwise Spearman’s rank correlation coefficient (L) was calculated using the score vectors and converted into pairwise distances (1-L). These distances were then used to cluster the genes into functionally related modules, constrained to a maximum of twenty genes, using agglomerative hierarchical clustering. Genes within a module exhibit the strongest topological similarities to each other within the multiplex network. Research team members were then assigned modules to further biologically characterize, enabling parallelized interpretation that was then united into a single conceptual model.

### Network layer ablation

The relative contribution of a network layer to the overall MENTOR clustering was tested by ablating the layer and then comparing differences in module membership of each gene relative to the MENTOR module assignments when using all layers. Set differences were calculated by using the adjusted Rand index to compare the clustering between the original 10-layer multiplex to each iteration of a 9-layer multiplex with one ablated network layer.

### Drug repurposing

Using DrugBank ^144^, we identified which of the 194 GRIN-retained genes were known drug targets. Drugs were annotated with the following categories: approved (by FDA/EMA/regulatory authorities), inducer of gene (including if known to be clinically significant), inhibitor of gene (including if clinically significant), agonist of gene, antagonist of gene, experimental use of drug, investigational use of drug, and/or nutraceutical use of drug. Drugs were also annotated if they were a known HIV ART based on one of the following classes: CCR5 inhibitors, fusion inhibitors, integrase inhibitors, nucleoside reverse transcriptase inhibitors (NRTIs), non-nucleoside reverse transcriptase inhibitors (NNRTIs), pharmacokinetic enhancers, or protease inhibitors.

## Data Availability

All data produced in the present study are available upon reasonable request to the authors.

## Acknowledgements

This work was supported by NIH grants P01 AA029545 (KX, DAJ), U24 AA020794 (KX), U10 AA013566 (KX), R01 DA051908 (DAJ, EOJ), and R01 DA051913 (DBH, DAJ). This research used resources of the Oak Ridge Leadership Computing Facility at the Oak Ridge National Laboratory, which is supported by the Office of Science of the U.S. Department of Energy under Contract No. DE-AC05-00OR22725. The Veterans Aging Cohort Study is funded by grants P01 AA029545, U24 AA020794, and U10 AA013566. The authors appreciate the support of the Veterans Aging Study Cohort Biomarker Core and the contributions of the VACS participants. This study was approved by the Institutional Review Boards at all VACS locations, which includes written consent before enrollment. The views and opinions expressed in this manuscript are those of the authors and do not necessarily represent those of the Department of Veterans Affairs or the United States government. This work uses data provided by patients and collected by the VA as part of their care and support.

## Author contributions

Conceptualized project: DAJ, EOJ

Provided funding: KX, DBH, DAJ, EOJ

Oversaw analysis/experiments: DAJ, EOJ

Performed analysis/experiments: KAS, MSM, XZ, WC, BCQ, CW, AT, PK, ML, RM, KX, BEA, DBH, DAJ, EOJ

Writing and revising of the paper: KAS, MSM, BCQ, DBH

## Ethics declarations

The authors declare no competing interests.

---

**Supplementary Figure 1.**
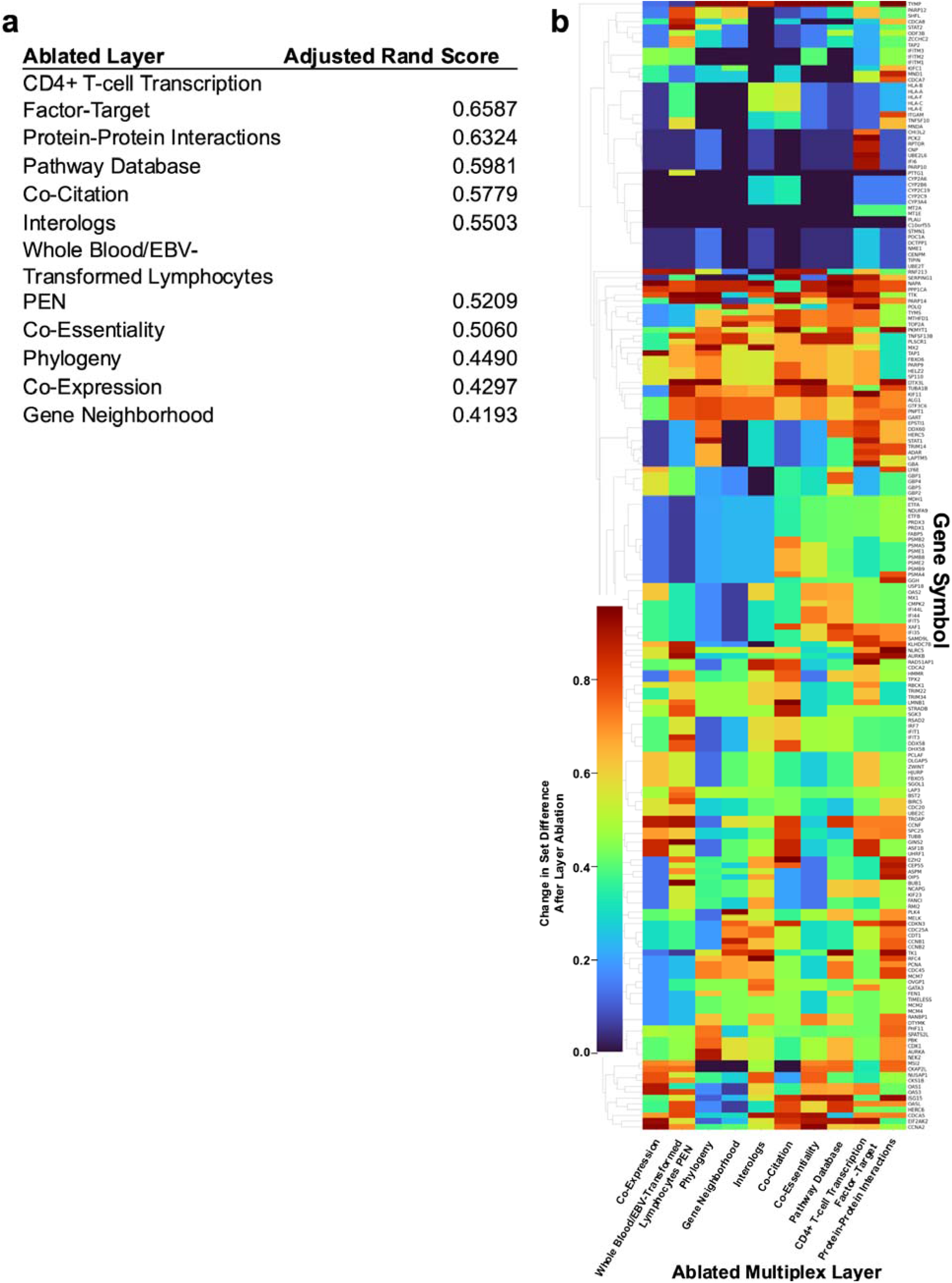
Multiple lines of evidence are necessary for MENTOR module interpretation. **a.** Running MENTOR with individual layers ablated resulted in different gene clusterings as measured by adjusted Rand score. Removing CD4+ T-cell transcription factor-target relationships resulted in the biggest disruption of gene membership within each dendrogram module. **b.** Gene-level visualization of module membership changes caused by ablating individual lines of biological evidence.

